# Feeding and swallowing outcomes of children receiving long-term ventilation: A scoping review protocol

**DOI:** 10.1101/2023.06.16.23291509

**Authors:** Sabrena Lee, Jeanne Marshall, Michael Clarke, Christina H Smith

## Abstract

**Background:** The last three decades have seen a growth in the number of children requiring long-term ventilation. Children with long-term ventilation present with underlying respiratory and neurological conditions that place them at risk of feeding and swallowing difficulties. To date, a scoping review or systematic review investigating the feeding and swallowing outcomes of children with long-term ventilation needs has not been conducted.

**Aims:** This paper describes a protocol for a scoping review of the feeding and swallowing outcomes of children receiving long-term ventilation.

**Methods:** This scoping review protocol will utilize the Joanna Briggs Institute scoping review methodology guideline. Our review will focus on the feeding and swallowing outcomes of children aged 0 to 18 years with long-term ventilation needs. A full search strategy initially created by the authors and a research librarian was conducted on the PubMed database. Following this, pilot testing took place to determine discrepancies in eligibility criteria. A full search strategy will be conducted across several databases. A data extraction form has been developed by the authors and will be used during the scoping review process.

**Discussion:** This protocol has been created to provide a rigorous and comprehensive basis for undertaking a scoping review. All necessary steps have been completed in order to commence the scoping review.

**Registration:** This scoping review protocol was registered on Open Science Framework on the 26^th^ November 2021 (Registration DOI 10.17605/OSF.IO/NQBPD)

## 1. Background

The aim of this paper is to present a protocol for a scoping review investigating the feeding and swallowing outcomes of children who require long-term ventilation.

### 1.1 Long-Term Ventilation in the Paediatric Population

Long-term ventilation (LTV) is defined as the dependence on various types of respiratory support for longer than three months from initiation of support and once an individual is medically stable (1). LTV includes continuous positive airway pressure ventilation (CPAP) and bilevel positive airway pressure ventilation (BiPAP) (2). CPAP delivers one constant pressure to stent the airways open and improve oxygenation in children with obstructive airway conditions, such as obstructive sleep apnea (3). BiPAP provides additional respiratory support by delivering different pressures during inspiration and expiration. It can also be used to maintain a child’s breathing rate and aid in the removal of excessive carbon dioxide in the lungs. BiPAP is therefore predominantly used to reduce respiratory effort and improve ventilation in children with centrally-driven respiratory conditions, such as those who do not take adequate breaths or those who have muscle weakness and are not able to take large enough breaths (3).

Ventilation can be delivered non-invasively (via a nasal or face mask) or invasively (via an artificial airway, such as a tracheostomy tube) (2,4). The ventilatory support received can also vary in the duration it is used over a 24-hour period (with increased duration of use typically indicative of increased severity of respiratory disease) (2). It is important to note that the use of high flow oxygen is not included as a form of LTV in the United Kingdom (UK) and therefore will not be included in this scoping review (National Confidential Enquiry into Patient Outcome and Death, 2020).

Indications for LTV include a wide range of sleep and respiratory disorders that fall into three main categories: a) chronic respiratory disease, b) respiratory muscle weakness, and c) central nervous system impairment (5). Children with LTV commonly have underlying medical conditions including chronic neonatal lung disease, cystic fibrosis, musculoskeletal weakness, airway malacia, congenital airway and lung abnormalities, and sleep disorders associated with impairment in neurological function and central respiratory drive (6).

The number of children with LTV needs have increased ten-fold in the last 15 years (5). It is hypothesized that this is due to the rising number of children living with complex chronic conditions and advances in medical technologies that have provided for use of ventilatory devices at home (7). In the UK, more than 3061 children and young people required LTV between the years of 2016 and 2018 (1). As this life-saving technology has allowed new possibilities of survival for children with complex and chronic medical needs, the number of children receiving LTV is anticipated to rise in the future (1). Therefore, prioritizing research in this area is vital for optimizing both current and future care for these patients and their families.

### 1.2 Feeding and Swallowing Difficulties in the Paediatric Population

Feeding is a broad term that encompasses any of the steps involved in eating and drinking (8). This includes a range of processes including bringing food and drink to the mouth, manipulating the bolus in the oral cavity, and swallowing it into the oesophagus. From birth, the development of feeding occurs in line with overall neurodevelopment and psychosocial skills (9). Typical feeding milestones begin with breast and bottle feeding as a newborn, to the introduction of solids at four to six months of age, and gradual transition through food textures and mastery of drinking skills throughout childhood (9). It has been widely accepted that competence in feeding is attained by approximately three years of age, although recent research suggests that the development of feeding skills may continue into the school-age years (10). Age-appropriate feeding development requires a complex interaction between the cognitive, sensorimotor, musculoskeletal, cardiopulmonary, and digestive systems (11). Therefore, disruptions in medical status, nutrition, feeding experiences, and the environment can have a significant impact on a child’s oral feeding journey and result in delayed or impaired feeding (11).

Swallowing is an integral part of feeding and involves the transport of liquid and food from the mouth to the stomach (8). Dysphagia is an impairment in swallowing and can occur in children with feeding difficulties (12). Dysphagia can lead to aspiration - the entry of fluid or food into the airway during swallowing, which poses a risk to a child’s respiratory health (9). Over time, dysphagia can lead to negative health consequences including aspiration pneumonia, failure to thrive, malnutrition, and dehydration (12).

Children with feeding and swallowing difficulties frequently present with one or more of the following characteristics: (a) difficulty with age-appropriate foods or fluids, (b) refusing an appropriate variety or amount of food or fluids, (c) undesirable mealtime behaviours, (d) difficulty with self-feeding skills, (e) difficulty using feeding devices and utensils and (f) displaying issues with growth (13). Feeding difficulties may or may not involve difficulties in swallowing (13). The term Paediatric Feeding Disorder (PFD) has recently been proposed as an umbrella term to capture the multifaceted and complex nature of paediatric feeding difficulties. PFD is defined as “impaired oral intake that is not age-appropriate, and is associated with medical, nutritional, feeding skill, and/or psychosocial dysfunction” (Goday et al., 2019, p. 124).

### 1.3 Feeding and Swallowing in Children with LTV Needs

The clinical presentation of feeding and swallowing in the child with LTV needs is frequently complex and multifactorial. Commonly, in a clinical setting, these children present with primary dysphagia secondary to their respiratory needs and/or other medical comorbidities (14). This group may also, however, experience chronic feeding difficulties in the longer term due to limited exposure to developmentally appropriate feeding experiences that, in turn, result in oral aversion and learnt refusal behaviours (15). The true nature and characterization of PFD in this cohort of children has not been formally explored in a research setting.

#### 1.3.1 Acute Feeding and Swallowing Difficulties

LTV is commonly prescribed for children with chronic respiratory disease, respiratory muscle weakness, and central nervous system impairment (5). From birth, successful oral feeding requires synchrony of the oral, pharyngeal, and respiratory mechanisms (11). This can be impeded by any medical condition that directly or indirectly impacts on these systems (11). Children who require LTV frequently present with underlying respiratory issues that commonly impact on safe coordination of breathing while eating and drinking. As respiration and swallowing share common structures in the pharynx, an impairment in one system can significantly affect the other (14). Indeed, dysphagia associated with respiratory morbidity has been well documented in the preterm population (Dodrill, 2011). Dysphagia is also prevalent in children who present with other diseases of the airways and lungs, airway malformations, and craniofacial malformations (14).

The neural control of swallowing lies within the cerebral, midbrain, and brainstem regions of the central nervous system, and therefore specific impairments in neurological function can have drastic effects on oral feeding (17). LTV is commonly used to support children with conditions affecting their central respiratory drive or neurological function, such as cerebral palsy, acquired/traumatic brain injury, and neuromuscular conditions (6). These populations are also at risk of dysphagia (14). An association between length of ventilation and dysphagia has been seen in children with traumatic brain injury (18).

The direct impact of the presence of LTV in the upper airways on swallow function is continuing to be researched in the field of paediatric feeding and dysphagia (Hirst et al., 2017). Children with invasive LTV via tracheostomy may experience changes in swallowing physiology due to the presence of the tracheostomy tube causing altered subglottic pressure, reduced sensation, and reduced movement of the larynx (12). It has also been suggested that the positive pressure delivered directly into the airways during non-invasive ventilation may alter airflow pressures and physiology, thus impeding closure of the airway during the swallowing process (20,21). However, this is currently an evolving area of research.

#### 1.3.2 Persisting Feeding and Swallowing Difficulties

Any medical illness, injury, or developmental delay that impacts on a child’s feeding development may lead to persisting feeding difficulties (11). Concerns regarding swallowing safety or medical instability at the time of initiation of LTV may result in the need for non-oral feeding, which can significantly disrupt the infant’s oral feeding journey (22). Indeed, a large majority of children with LTV via tracheostomy are fed via nasogastric tube in the short-term and gastrostomy tube in the long-term (23). Reduced oral stimulation, especially during critical periods of feeding development, may result in dysfunctional feeding patterns such as oral aversion and feeding refusal (24). In addition, the presence of invasive medical interventions, such as respiratory support and feeding tubes around the face and mouth, can result in negative experiences and oral hypersensitivity that lead to subsequent feeding problems (Burklow et al., 2002; Dodrill et al., 2004). These factors have been associated with slower transition from non-oral to oral feeding methods within the acute setting (26).

An association between oral feeding disorders and history of ventilation, aspiration and tube feeding has been reported (27). Duration of invasive ventilation has been identified as a risk factor for the development of eating problems in very low birth weight children (28). Similarly, prolonged length of ventilation has also been associated with interruption of exclusive breastfeeding in premature infants (29). An increased risk of dysfunctional feeding behaviours has been associated with the number of days of mechanical ventilation in premature children (30).

While it can be difficult to differentiate the distinct aetiology of the presenting feeding difficulties in children with LTV, it is important to consider both swallowing and feeding difficulties in a holistic manner that accurately reflects the complexity of paediatric feeding disorder within this population.

### 1.4 Reviews to date

A preliminary search of Cochrane Database of Systematic Reviews, JBI Evidence Synthesis, Open Science Framework, Figshare, ERIC (ProQuest), PubMed and CINAHL Plus was conducted in October 2021.

To date, no scoping or systematic reviews investigating the feeding outcomes of children with LTV have been identified. However, scoping reviews exploring the feeding outcomes of particular cohorts that include some children with LTV were found. Fucile et al. (2021) completed a scoping review to investigate the risk factors associated with long-term feeding problems in preterm infants. The duration of mechanical ventilation in neonatal intensive care was found to be a risk factor for long-term feeding problems in the preterm population (22). Pados et al. (2021) conducted a meta-analysis of the prevalence of problematic feeding in young children who were born preterm. Results revealed that long-term feeding difficulties are highly prevalent in preterm children from birth to four years of age, however, most research studies in this review excluded infants with the highest risk for problematic feeding including chronic lung disease and need for respiratory support (31).

### 1.5 Review objective

The research topic of feeding and swallowing outcomes of children with LTV has not been comprehensively explored previously. The current scoping review will aim to map the breadth of literature in this field of research by collating sources of evidence from a variety of global multidisciplinary areas. As the current research in this specific topic may be limited, a scoping review will allow all relevant forms of literature, including grey literature, to be used to inform the review question. This will allow the authors to systematically summarise the available evidence and in turn, identify existing gaps in the literature to determine potential directions for future research.

## 2. Review question

The primary review questions are (a) ‘What specific swallowing and feeding characteristics do children with LTV needs present with?’ and (b) ‘’What impacts do these swallowing and feeding difficulties have on health status and quality of life?’

## 3. Keywords

Children; long-term ventilation; feeding; swallowing; scoping review

## 4. Eligibility Criteria

### 4.1 Participants

The target population are children (aged 0-18 years). This includes children who were born preterm (before 40 weeks of full-term pregnancy). Children with all medical backgrounds will be included. Studies that include animal experiments and/or observations will be excluded.

### 4.2 Concept

This scoping review will include any literature that describe outcomes regarding feeding and/or swallowing in infants and children who have received LTV. Studies regarding eating disorders (e.g., anorexia nervosa, bulimia nervosa, binge eating disorder) will not be included.

### 4.3 Context

This scoping review will report on research literature describing LTV. The definition of LTV includes the use of CPAP or BiPAP in a non-invasive or invasive method for at least three months from initiation of support and once medically stable (4). Studies that include children who require LTV for varying durations of time throughout the day and night will be accepted. Studies that do not include children with LTV (for example, in cases where ventilation was provided for less than three months duration) will be excluded.

International research will be included in the scoping review. This scoping review will not have any other exclusion criteria regarding demographic, cultural or geographical-based contexts.

### 4.4 Types of Sources

Due to the heterogenous nature of available sources in this field, this scoping review will include all types of literature including scoping reviews, systematic reviews, meta-analyses, guidelines, policy documents, websites, letters, articles, and research papers. Both qualitative and quantitative research studies will be included to address the review question comprehensively. Posters and conference abstracts will not be included as these are unlikely to contain sufficient information to answer the review questions. Literature that is not available in English will also be excluded. The inclusion criteria reflect the Joanna Briggs Institute (JBI) methodology for conducting scoping reviews (32).

## 5. Methods

This protocol has been completed using the Joanna Briggs Institute (JBI) SUMARI protocol template (32). The JBI scoping review methodology, with inclusion of the PRISMA-ScR reporting guideline, will be used to conduct this scoping review (33).

### 5.1 Search Strategy

In accordance with JBI review methodology, a three-step search strategy will be employed. Firstly, an initial limited search of all relevant online databases, including PubMed, MEDLINE (Ovid Version), ERIC (ProQuest), and CINAHL Plus, will be conducted. The title and abstract of retrieved papers will be analysed in order to identify keywords and index terms. Secondly, all identified keywords and index terms will be used to perform another search across the same bibliographic databases. Thirdly, a search of relevant literature within the reference list of all included documents will be conducted.

If required, reviewers will contact authors of retrieved papers to clarify information necessary to answer the scoping review question. The entire search process will be carried out by the first author, in collaboration with the co-authors, as well as a research librarian.

Studies published in English (and already translated into English by the publishing journal) will be included. Studies published since 2000 will be included, as the research field of LTV has significantly developed within this time frame (7).

The search strategy will aim to contain both published and unpublished literature extending the breadth of the review as much as possible. Therefore, in order to source appropriate policies and guidelines, an additional search for grey literature using handsearching via a generic search engine in Google will be conducted. The JBI guideline will be used to ensure this is completed in a systematic manner.

Please see Appendix I for the full search strategy used in one database (PubMed). The scoping review will contain evidence of the full search strategy from all databases included.

### 5.2 Source of Evidence Selection

Pilot testing of source selectors will be conducted prior to evidence selection. Pilot testing will consist of the research team screening a random sample of 25 journal titles/abstracts using the eligibility criteria and definitions document. Two authors will determine discrepancies present and adjust eligibility criteria and definitions if required. Once 75% agreement is achieved, the evidence selection process will then follow.

Once the full search strategy has been completed across all databases, identified citations will be collated and uploaded into Endnote Version 20.2 2013 (Clarivate). This database will then be imported into the online systematic review software, Covidence. Duplicates of citations will be removed before title and abstract screening is commenced by two authors. Following this, the full texts of relevant articles will be retrieved and assessed by the same two authors. Any disagreements that arise during this process will be resolved through discussion with the research team. This process will also be repeated for a search of relevant grey literature. Full texts will be assessed by two authors according to the inclusion/exclusion criteria, and source information will be managed on an electronic database. The results of this evidence selection process will be reported in the final scoping review in a narrative description and presented in a PRISMA-ScR flow diagram as per JBI Methodology guidelines.

### 5.3 Data Extraction

Data extraction will be guided by a fit-for-purpose data extraction form developed by the research team that includes details about the participants, concept, context, methodology, and key findings in the context of the primary and secondary review questions. This data extraction form is presented in Appendix II.

Prior to full data extraction, two authors will independently pilot the data extraction form on at least three sources of evidence to ensure that all relevant results are extracted. This is in accordance with the pilot testing procedure suggested in the JBI methodology guidelines. Following this, all data will be extracted from all papers included in the scoping review by one reviewer. Any modifications to the proposed data extraction tool during the scoping review process will be documented and presented in the final scoping review. The final data set will be reviewed and discussed by the team to come to consensus with regards to findings and implications.

### 5.4 Data Analysis and Presentation

As per the JBI Methodology guidance, this scoping review will analyse and extract data in a descriptive manner, and present results in both narrative and tabular forms. Information regarding evidence selection will be presented in a flowchart to demonstrate the full search strategy and process of inclusion of evidence. Due to the wide range of feeding outcomes that may be described, scoping review results may be presented in multiple forms to ease reader viewing and understanding of information. This will be useful in identifying and summarizing evidence on the review question, as well as highlighting research gaps in the topic. As per the data extraction form, information will be categorized by details related to the review question.

## 6. Conclusions

The increasing number of children with long-term ventilation needs in recent years has led to a growing urgency to plan and deliver holistic health services that meet their needs (1). The known risk factors for feeding and swallowing disorders in this population mandate further exploration and characterization of these outcomes in this group, as a fundamental part of this process. This a priori protocol has described the framework that will be utilized when conducting a scoping review to explore this research topic. The objectives, design, methods, analysis, and presentation of results have been outlined, which allows for transparency of the scoping review process. A complete scoping review will follow, as per the framework set out in this protocol, and results will be disseminated and presented as planned.

## Data Availability

All relevant data are within the manuscript and its Supporting Information files.

## Acknowledgements

The research team would like to acknowledge the contribution of the research librarians at the University College London Language and Speech Science Library who provided valuable assistance during this project.

---

**Appendix I:**
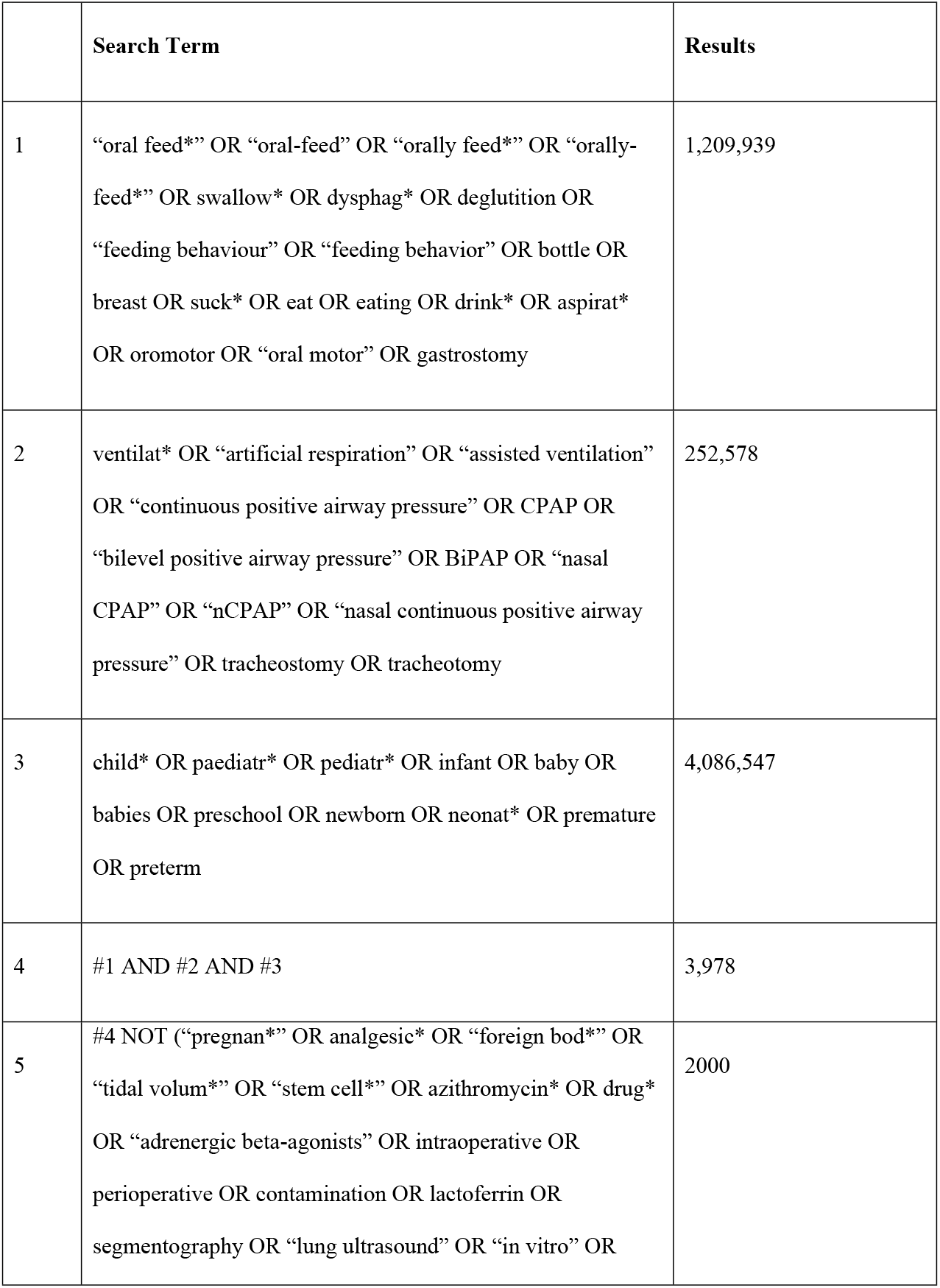
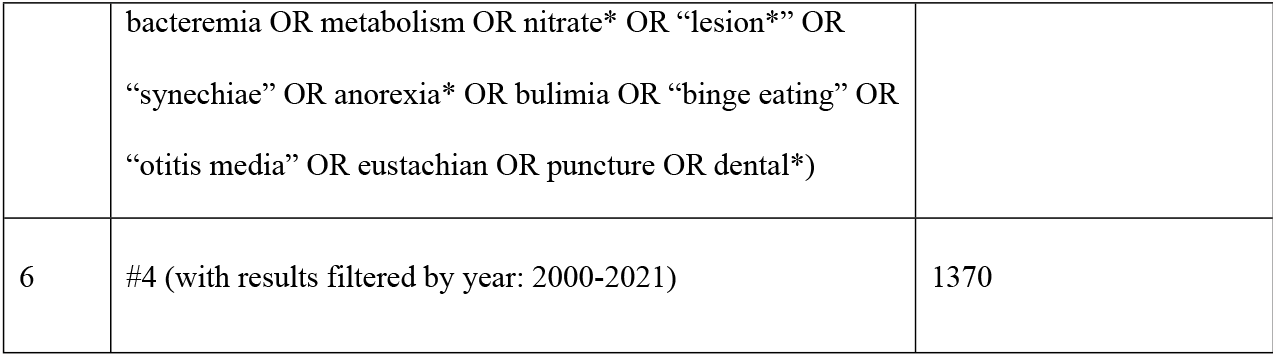
Full search strategy for PubMed. **Date of search: 13^th^ December 2021**

**Appendix II:**
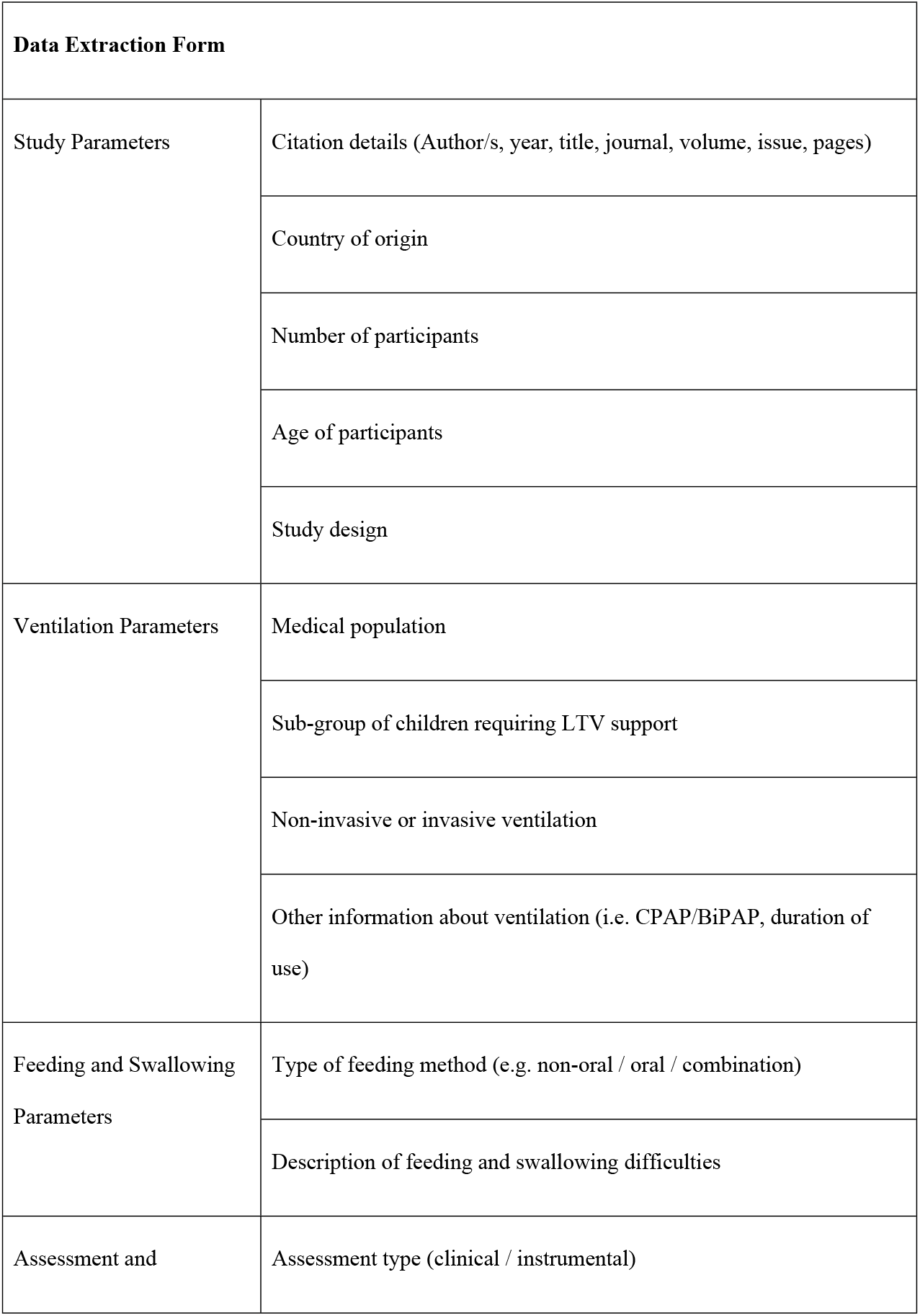
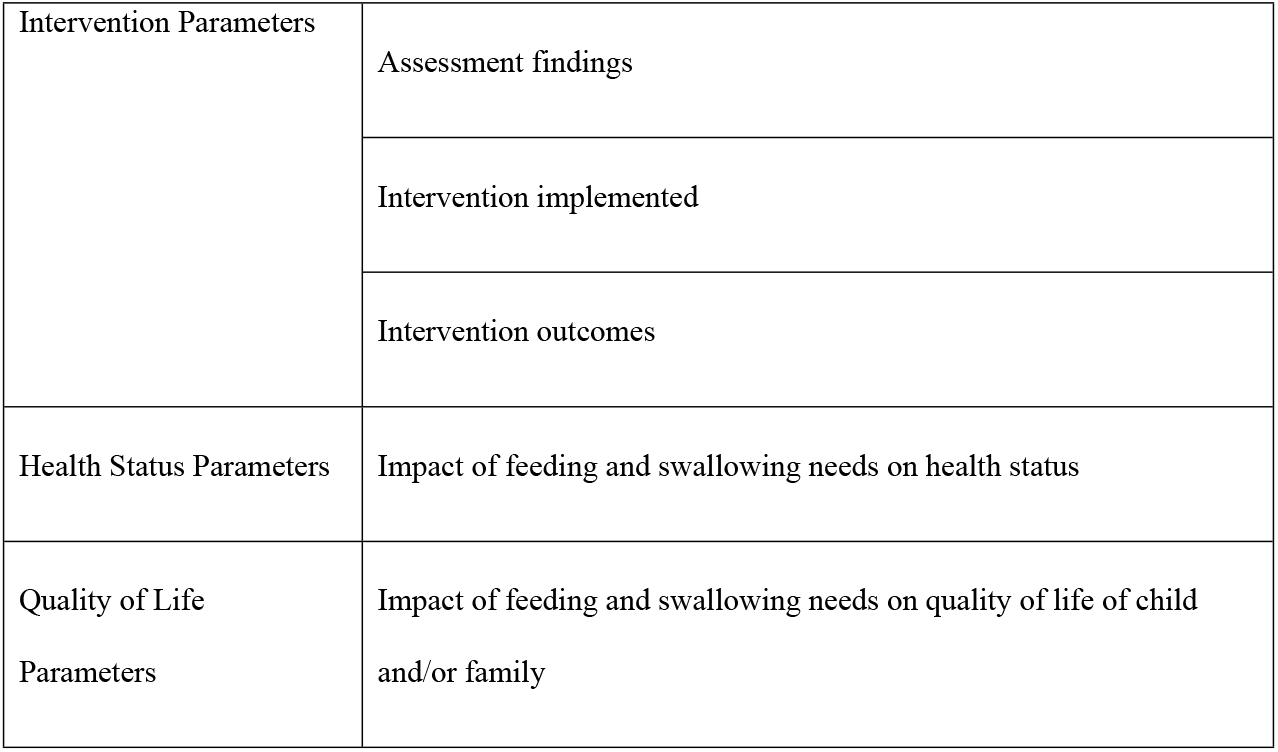
Data Extraction Form.

